# Carbapenemase-producing Enterobacterales colonisation status does not lead to more frequent admissions: a linked patient study

**DOI:** 10.1101/2024.01.09.24300945

**Authors:** Michael J Lydeamore, Tjibbe Donker, David Wu, Claire Gorrie, Annabelle Turner, Marion Easton, Daneeta Hennessey, Nic Geard, Ben P. Howden, Ben S. Cooper, Andrew Wilson, Anton Y Peleg, Andrew J Stewardson

## Abstract

**Background:** Hospitals in any given region can be considered as part of a network, where facilities are connected to one another – and hospital pathogens potentially spread – through the movement of patients between them. We sought to describe the hospital admission patterns of patients known to be colonised with carbapenemase-producing Enterobacterales (CPE), and compare them with CPE-negative patients.

**Methods:** We performed a linkage study in Victoria, Australia, including datasets with notifiable diseases (CPE notifications) and hospital admissions (admission dates and diagnostic codes) for the period 2011 to 2020. Where the CPE notification date occurred during a hospital admission for the same patient, we identified this as the ‘index admission’. We determined the number of distinct health services each patient was admitted to, and time to first admission to a different health service. We compared CPE-positive patients with four cohorts of CPE-negative patients, sampled based on different matching criteria.

**Results:** Of 528 unique patients who had CPE detected during a hospital admission, 222 (42%) were subsequently admitted to a different health service during the study period. Among these patients, CPE diagnosis tended to occur during admission to a metropolitan public hospital (86%, 190/222), whereas there was a greater number of metropolitan private (23%, 52/222) and regional public (18%, 39/222) hospitals for the subsequent admission. Median time to next admission was 4 days (IQR, 0-75 days). Admission patterns for CPE-positive patients was similar to the cohort of CPE-negative patients matched on index admission, time period, and age-adjusted Charlson comorbidity index.

**Conclusions:** Movement of CPE-positive patients between health services is not a rare event. While the most common movement is from one public metropolitan health service to another, there is also a trend for movement from metropolitan public hospitals into private and regional hospitals. CPE-positivity does not appear to impact on hospital admission frequency and timing. These findings support the potential utility of a centralised notification and outbreak management system for CPE positive patients.

## Introduction

Carbapenemase-producing Enterbacterales (CPE) infections are a significant public health concern [1]. In Victoria, Australia, CPE infections have been steadily increasing in recent years despite a centralised ‘search and contain’ policy [2], with acquisition primarily associated with international travel and healthcare exposure [2]–[4]. This trend poses a serious threat to patient outcomes, as CPE infections are associated with increased morbidity, mortality, and healthcare costs [5].

Hospitals in any given administrative or geographical region can be considered as part of a network, where facilities are connected to one another through the movement of patients between them [6][7]. Patients may either be directly transferred from one hospital to another, or admitted to a given hospital at some time after discharge from another hospital. In this way, patients colonised by a multidrug resistant bacteria, such as CPE, in one hospital can transmit it to patients in another [8]–[10] with models suggesting a region-wide approach could be necessary to achieve control of infection [11]–[13].

This inter-hospital transmission risk is acknowledged in the Victorian Department of Health’s CPE guidelines [14]. Victoria has no centralised laboratory information or alert system to share information about CPE colonisation; therefore, patients colonised by CPE, and their primary healthcare providers, are instructed that in case of future hospital admission, they should inform the facility about their CPE colonisation status to facilitate isolation with appropriate contact precautions. There is currently no data about the frequency with which patients colonised by CPE present to other healthcare facilities. Such data would help quantify the transmission risk posed by patients colonised with CPE, particularly in the context of reliance on self-reported CPE-colonisation status.

Our primary aim was to describe the hospital admission patterns of patients known to be colonised with CPE, using linked patient data, including the type of health service and timing of admissions. Our secondary aim was test the hypothesis that patients colonised with CPE have different admission patterns to other healthcare system patients.

## Methods

### Setting and population

Victoria, Australia, has a population of 6.5 million people, with a median age of 38 years [15]. The hospital system consists of more than 300 hospital campuses, which vary greatly in size, case mix and services.

Hospitals in Victoria are classified as “public” or “private” services. Public health services are freely available to the entire population, with funding provided by the state and federal government. Private health services service a subset of patients, generally those with private health insurance.

Public health services usually govern several hospital campuses, with 149 public hospital campuses divided into 80 unique health services. Private hospitals have a much more complex and varied governance structure, with dynamic ownership structures. The hospital system in Victoria operates using a ‘devolved governance’ structure [16], with each health service having its own board that dictates it’s policies.

The Victorian Guideline on Carbapenemase-producing *Enterobacteriaceae* for Health Services was released in December 2015, and involved a centralised genomic and epidemiological surveillance and response program [2]. CPE became a notifiable condition in Victoria in December 2018.

The study period was 1 January 2011 and 30 November 2020. All Victorian hospital admissions during the study period are included. Patients were identified as CPE-positive based on the presence of a notification in the Public Health Events Surveillance System (PHESS) database between May 2013 (the first notified case) and 30 November 2020 (N = 628). Patients without a recorded notification in this database were considered CPE negative.

### Data sources

The PHESS database contains patient demographics and epidemiologic data relevant to each case. For our analysis, we focused on the ‘calculated onset date’, determined as follows: if the patient’s acquisition date (that is, the date of transmission from another case) was known, it was used as the calculated onset date; if the acquisition date was unavailable, the specimen collection date was used instead; and if no other information was available, the calculated onset date was defined as the date when the notification was received by the health department. For the purposes of this analysis, calculated onset date is considered to be the best estimate of the date at which a patient became CPE positive.

The hospital admissions are sourced from the Victorian Admitted Episode Dataset (VAED) [17]. Linkage between the PHESS and VAED datasets was performed by the Centre for Victorian Data Linkage, as part of the Victorian Linkage Map and Integrated Data Resource. The linkage allowed build a sequence of admission and CPE status for each patient, and analyse how their admission patterns may have changed post-CPE positivity, and to compare population groups post-hoc.

### Analysis

Our hypothesis that CPE positive patients have similar healthcare-seeking behaviours to CPE negative patients is tested by analysing differences in two key metrics between CPE positive and CPE negative patients: the number of health services visited after diagnosis, and the time until the next admission after diagnosis.

To test this hypothesis, we use survival analysis to analyse the differences in estimated times of re-admission. The survival function, denoted *Ŝ*(*t*), represents the probability that the event of interest (here re-admission) has not yet occurred by time *t*. Specifically, we calculate Kaplan-Meier survival curves, calculated as

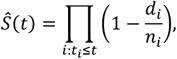

where *d*_*i*_ is the number of events that have occurred up to time *t*_*i*_ and *n*_*i*_ is the number of individuals who have not yet experienced the event or been censored up to time *t*_*i*_.

The admission associated with CPE identification (index admission) is chosen to be the admission which encloses the calculated onset date from the patient in PHESS. Index hospital admissions could be identified for 528 patients. Patients who were not admitted at the CPE onset date were removed from further analysis (N=100). Patients who died during the index admission were removed from further analysis (N = 81).

To determine the time of risk and the time of readmission event for conducting the survival analysis, the period of risk (time 0) was defined as the end date of the index admission. If the patient was transferred directly to another hospital campus within the same health service, then time 0 was the date of discharge from the health service. The event time was set as the beginning of the subsequent admission to a different health service. Any admissions occurring after time 0 to the same health service where the patient was initially diagnosed were excluded from consideration. This exclusion assumed that the assumption that the health service would have recorded the CPE positive status of patients and, as a result, would have taken proactive measures such as isolation. Patients who do not appear in an admission to a different health service to that of diagnosis, they are assumed to be censored at the end of the data period (30 November, 2020).

Four CPE-negative patient cohorts were created by sampling from the whole population using different matching criteria. These matching criteria were chosen to produce cohorts with increasing levels of similarity to the CPE-positive cohort:

- **Cohort 1. Random subset**: patients are randomly sampled from the population.
- **Cohort 2. Campus and time**: Patients are matched to CPE-positive patients based on the hospital campus and the quarter-year (i.e. 3-month period) of admission.
- **Cohort 3. Campus, time and age**: As per Cohort 2, plus the inclusion of five-year age band.
- **Cohort 4. Campus, time and comorbidities**: As per Cohort 2, plus the inclusion of age-adjusted Charlson Comorbidity Index category.

ICD-10-AM codes were used to calculate the age-adjusted Charlson Comorbidity Index [18]. Age adjustment was applied by increasing the Charlson score by 1 for every 10 years of patient age beyond 40 [19]. The resulting age-adjusted scores were categorized into four groups: None (0 points), Mild (1-2 points), Moderate (3-4 points), or Severe (>=5 points). To ensure robustness, the sampling process was repeated 100 times with replacement. Each CPE-positive patient had at least 10 potential matching CPE-negative patients identified for the analysis.

Kaplan-Meier survival curves were calculated separately for each patient cohort, using R version 4.1.0 and the survival package.

### Ethics

Ethics permission was obtained from the Alfred Health ethics board, submission number 445/21.

## Results

The study population includes 528 unique CPE positive patients. Figure 1 shows the number of notifications for confirmed cases, by month, over the data period. We note that CPE was made notifiable in Victoria in December of 2018, with a increase in notifications from that time. The low notifications toward the end of 2020 are likely due to data censoring at the end of the study period. Patients can have more than one notification for CPE i.e. if they are colonised by more than one CPE, with distinct species of Carbapenemase gene. All notifications are shown in Figure 1, but only the earliest onset is considered for further analysis.

**Figure 1:**
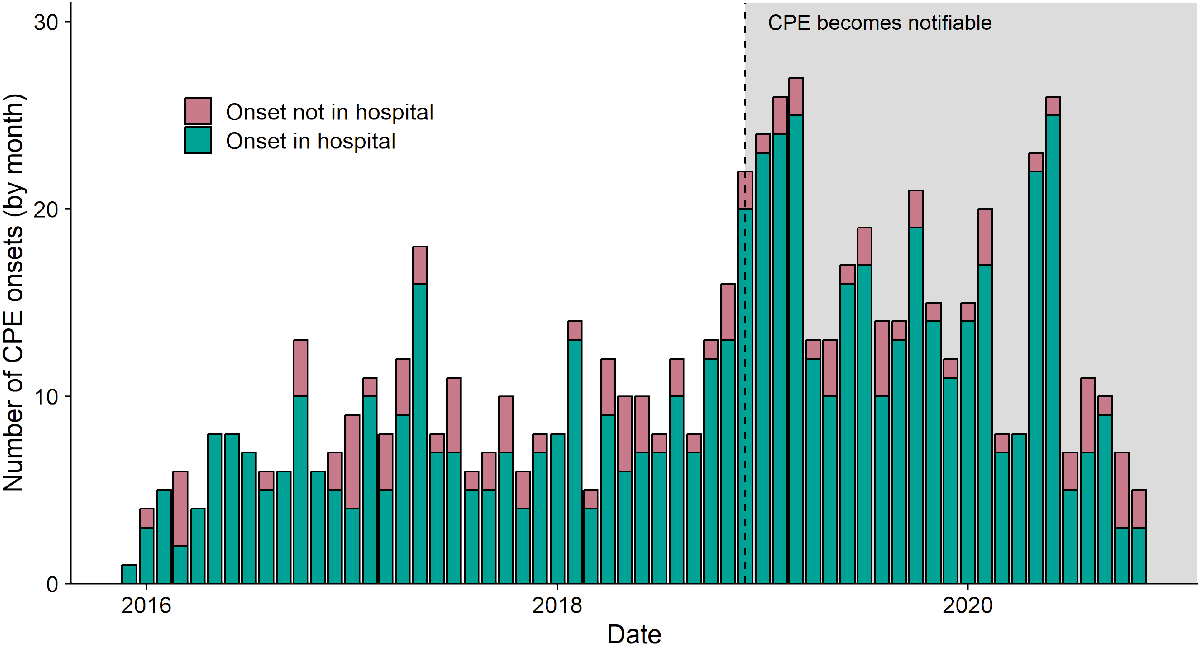
Number of CPE notifications, by onset date, grouped by month notified to the Victorian Department of Health. Grey shaded region indicates that CPE infection had become notifiable, although surveillance had been established in December 2015. Coloured bars represent whether the patient’s calculated onset date falls in a hospital admission. The proportion of onsets within and outside of a hospital admission remains approximately constant over time.

Patient demographic summaries are contained in Table 1. Of the 528 CPE positive patients included in the study, almost two thirds (63%) were male. The age distribution of positive patients tended toward older age groups, with 59% of patients being over 60 years of age.

**Table 1:** Patient demographics for CPE positive patients. Age-adjusted Charlson Comorbidity index is calculated on the earliest admission containing the earliest calculated onset date.

| Gender | N (%) | Median Age-adjusted Charlson Comorbidity Index [IQR] |
| --- | --- | --- |
| Male | 333 (63%) | 2 [0-5] |
| Female | 195 (37%) | 2 [0-4] |
| Age |  |  |
| 0-4 | 10 (2%) | 0 [0-2] |
| 5-19 | 8 (2%) | 0 [0-0.5] |
| 20-39 | 81 (15%) | 0 [0-2] |
| 40-59 | 117 (22%) | 2 [0-4] |
| 60-79 | 225 (43%) | 2 [1-5] |
| 80+ | 87 (16%) | 3 [1-6] |
| Acquisition location |  |  |
| Metro, private | 38 (7.3%) | 2 [0-5] |
| Metro, public | 472 (91%) | 2 [0-5] |
| Rural, public | 18 (3.4%) | 1 [0-4] |
| Rural, private | 0 (0%) | – |

Of the 528 patients in our cohort, 222 (42%) were admitted to more than one health service within the study period. Among these patients who moved between health services, most (90.6%, 201/222) notifications occurred at times when patients were admitted into a public hospital (Figure 2). Of those diagnosed while in a public hospital, 84% (169/201) had their subsequent admission in a public hospital, with the remaining 16% being admitted to a private hospital. For those diagnosed in private health services (n=21), two thirds were re-admitted to a different health service. We note that a very large percentage of patients (306 [58%]) were never admitted to a different health service within the study period. These patients are not included in the description of diagnosis-readmission pathways.

**Figure 2:**
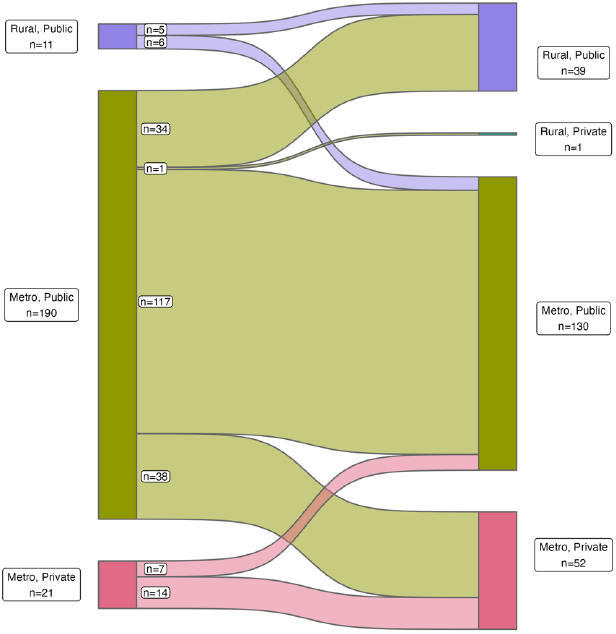
Sankey diagram showing the flow of patients from location of diagnosis admission to location of subsequent admission. Majority of patients are diagnosed in metropolitan, public hospitals, and are re-admitted there.

The number of unique health services visited after the beginning of risk period (including the index hospitalisation) is presented for CPE-positive patients and the four matched cohorts (Figure 3). While most CPE-positive patients are only admitted to one health service, 155 (29%), 44 (8.3%) and 20 (3.7%) patients are admitted to two, three, and four or more distinct health services, respectively. Certain factors, such as index hospital campus and time, were found to impact on the number of unique health services visited. However, it appears that severity of illness is the main driver for this measure of behaviour, rather than a specific CPE diagnosis.

**Figure 3:**
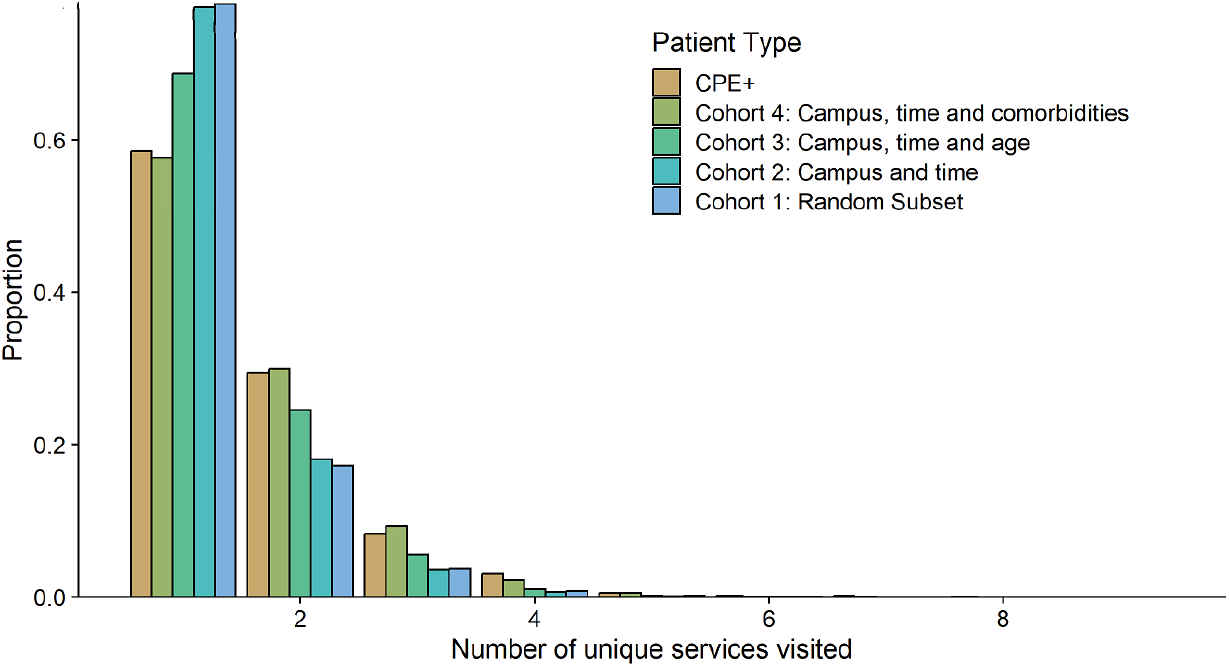
Number of unique health services visited for patients diagnosed with CPE, and the four matched cohorts. The service of admission was determined using the admission date, or the notification date for patients.

Time from the end of the index admission until next admission to a different health service is shown in shown for CPE positive patients, and the four matched cohorts (Figure 4). At 90-days, 1-year, and 2 years, 40%, 50% and 57% of CPE-positive patients had been admitted to a different health service. There is no difference in the median survival times (i.e. time until next admission) between CPE-positive patients and the cohort matched on Charlson Comorbidity Index bracket, which means that patients who are CPE-positive re-appear in hospital no more frequently than those who have not tested positive. There is a large proportion of censored patients in this dataset, both in the CPE-positive and matched cohorts, which is reflective of patients appearing in hospital once, and never returning.

**Figure 4:**
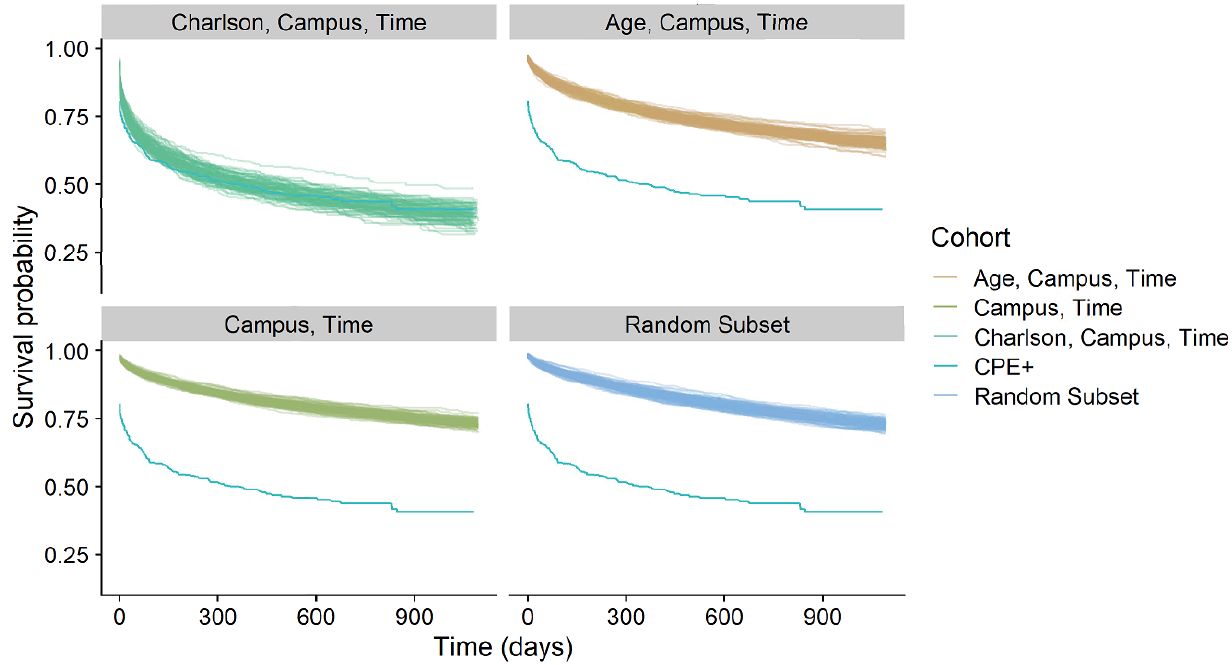
Survival curves for patients who have tested positive for CPE, against those who have not tested positive. Each line shows a survival curve for a particular sampled population. As matching criteria becomes stricter, the difference between CPE positive patients and the matched populations decreases.

Of note, 71 (15.8%) patients experience a transfer to another health service immediately after discharge (i.e., have a time of event of 0 days). This is approximately double that observed in the most closely matched cohort of CPE negative patients (cohort 4). Of those who are directly transferred, three are censored (as they are discharged on the last day of the dataset). There are 48 patients who travel between public health services, and 13 who are diagnosed in a public service, then re-appear in a private service.

## Discussion

Patients known to be colonised with CPE are frequently re-admitted to different health services to the one they are diagnosed in. However, using survival analysis, we have shown that this frequency of readmission is not significantly higher than other healthcare system patients, controlling for comorbidities.

We have also shown that majority of CPE colonisation identification and re-admission occurs in public, metropolitan hospitals in Victoria. This is perhaps correlated with hospital capability and centrality.

One unexpected trend observed in this data is the bias toward male patients. This trend hasn’t previously been reported, and further investigation as to whether this is a health service coding issue or an actual trend is warranted to further the epidemiologic understanding.

This work uses administrative data on patient admissions and separations across Victoria, which includes a large amount of quality assurance testing and input restrictions. By combining with notifiable disease information, which is legally required to be reported upon detection, this analysis uses very high quality input data. By using statistical linkage, we were able to follow patient journeys through the healthcare system, which is highly valuable when considering patient healthcare seeking behaviour.

Testing for CPE infection is based on highly varied screening procedures, with increased screening being performed generally in intensive care units, and after an outbreak is detected [20]. As a result, it is unlikely that detection of CPE infection is perfect. This means that potentially mild cases of CPE colonisation (which are still contagious) may not be included in this analysis, and these patients may have different healthcare seeking behaviours.

Most studies of CPE colonisation are prospective studies [21], [22] that are focussed in one facility or ward. By using routinely reported data, we are able to get a greater understanding of the breadth of patients who may be admitted while colonised with CPE, and also understand their future health seeking behaviour. However, a key limitation of retrospective studies is the inability to change sampling strategies, or investigate trends further than what is present in the data.

The linked nature of the data used in this study is another key strength. The impact of inter-facility patient transfer has been studied previously for CPE [13] and other hospital associated infections [21]. These studies have only been able to draw associations between hospital incidence and patient admissions, while this data allowed us to look *explicitly* at the patterns of those colonised with CPE.

We focussed on hospital-acquired CPE infection. While this is likely to be the primary source of infection, community and household based transmission has been quantified previously [24]. We note that almost 16% of the patients in the notifiable disease database do not have an associated hospital admission. Many of these are likely returning travellers, but it is possible that community transmission could be a contributor to overall disease burden.

While we have shown that patients colonised with CPE are frequently transferred around the hospital system, we have not evaluated the impact this may have on transmission. Dynamic models of disease and its control are required for this purpose, and are a direction of future work. We have also not been able to identify the frequency with which patients are appropriately placed in contact precautions after CPE diagnosis when admitted to a different health service.

Patients colonised with CPE are not fundamentally different in their healthcare seeking behaviour than those who are similarly ill. From an infection prevention point of view, this means that direct transfers of patients who are known to be positive is still high, giving ample opportunity for between-facility transmission shortly after detection. This supports the potential utility of a centralised alert system to identify CPE positive patients on hospital presentation.

## Data Availability

Data used in this work are available via Zenodo at https://zenodo.org/doi/10.5281/zenodo.10467482

## Acknowledgements

We would like to acknowledge the Victorian Department of Health as the source of VAED and PHESS datasets for this study, and the Centre for Victorian Data Linkage (Victorian Department of Health) for the provision of the data linkage.

The opinions expressed are those of the authors do not represent the views, nor the policy directions, of the Victorian Department of Health.

### Appendix

**Table 2:**
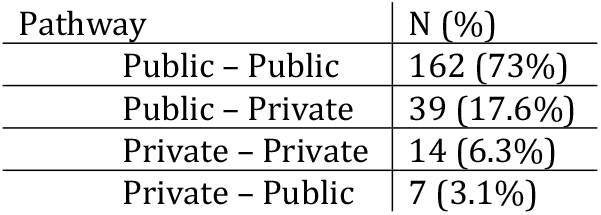
Number of re-admissions amongst non-censored patients who have tested positive for CPE, stratified by whether they were diagnosed and then visited public or private health services.

| Pathway | N (%) |
| --- | --- |
| Public – Public | 162 (73%) |
| Public – Private | 39 (17.6%) |
| Private – Private | 14 (6.3%) |
| Private – Public | 7 (3.1%) |

**Table 3:**
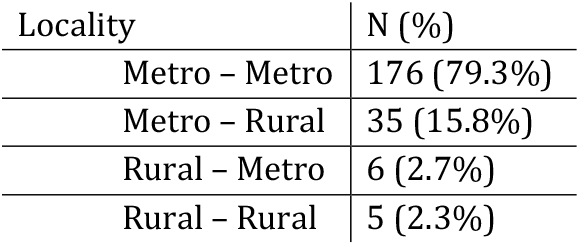
Number of re-admissions amongst non-censored patients who have tested positive for CPE, stratified by whether they were diagnosed and then visited metropolitan or rural health services. Metropolitan/rural classification follows the Department of Health region classifications.

| Locality | N (%) |
| --- | --- |
| Metro – Metro | 176 (79.3%) |
| Metro – Rural | 35 (15.8%) |
| Rural – Metro | 6 (2.7%) |
| Rural – Rural | 5 (2.3%) |

## Notes

### Competing Interest Statement

The authors have declared no competing interest.

### Funding Statement

This research was funded by the Australian National Health and Medical Research Council (GNT1156742). A.J.S. was supported by an Early Career Fellowship from the National Health and Medical Research Council (GNT1141398).

### Author Declarations

Ethics permission was obtained from the Alfred Health ethics board, submission number 445/21.

